# Speech Differences between Multiple System Atrophy and Parkinson’s Disease: a Multicenter Study

**DOI:** 10.1101/2024.02.23.24303241

**Authors:** Tom Hähnel, Anna Nemitz, Katja Schimming, Luise Berger, Annemarie Vogel, Doreen Gruber, Nils Schnalke, Stefan Bräuer, Björn H. Falkenburger, Florin Gandor

**Author notes:** **Corresponding author:** Dr. med. Tom Hähnel, Department of Neurology, University Hospital Carl Gustav Carus, Technische Universität Dresden, Fetscherstrasse 74, 01307, Dresden, Germany. No funding was obtained for conducting this study.

## Abstract

**Background:** Delineation of Parkinson’s disease (PD) from multiple system atrophy (MSA) can be challenging, especially in early disease stages, and clinical markers are needed for early detection of MSA. Speech characteristics have been studied as digital biomarkers in PD and ataxias, but there is only little data on MSA.

**Objectives:** To determine whether speech characteristics can serve as a biomarker to differentiate between MSA and PD.

**Methods:** 21 MSA patients and 23 PD patients underwent a battery of speech task assessments: text reading, sustained phonation and diadochokinetic tasks. Speech characteristics were extracted using the software Praat.

**Results:** MSA and PD speech can be described by the factors: “time and pauses”, “harsh voice”, and a factor containing “mixed speech characteristics”. After correcting for MDS-UPDRS III, four parameters and the “time and pause” factor showed significant differences between MSA and PD. MSA could be delineated from PD with Receiver Operator Characteristic Area Under the Curve (ROC-AUC) of 0.89 by a single speech parameter together with MDS-UPDRS III.

**Conclusion:** MSA can be differentiated from PD with good accuracy using only MDS-UPDRS III and one speech parameter as predictors. This outlines the importance of speech assessments to delineate MSA from PD to allow for differential diagnosis in movement disorders.

## Introduction

Multiple System Atrophy (MSA) is a progressive, neurodegenerative disease presenting with parkinsonism and/or cerebellar symptoms in combination with dysautonomia. Accuracy of MSA diagnosis is especially low at initial consultation and thereby delays or even prevents MSA patients from being enrolled into clinical trials that test for potentially disease modifying treatments in the early disease course (1).

Speech characteristics have been widely studied as a digital biomarker in PD (2) and in ataxias (3–7). In contrast, speech data in MSA is limited and only available from small cohorts (8–14). In addition, the majority of studies are from native Czech speakers. Therefore it is uncertain whether previous findings can be transferred to other languages (8,10,11,14). Thus, it remains unclear if speech characteristics can serve as a robust digital biomarker for differentiating between MSA and PD.

Speech in MSA is characterized by a mixed pathology of spastic, ataxic and hypokinetic speech symptoms (8,11,13,15). Because PD also has a high prevalence of hypokinetic speech (2), there is a considerable clinical overlap regarding speech impairment (11).

In contrast to PD, MSA shows a faster disease progression (16) and lacks levodopa responsiveness of Parkinsonian symptoms (17,18). Thus, it is characteristic that MSA patients have a higher motor impairment and disease burden with shorter disease duration compared to PD (16). Moreover, laryngopharyngeal dysfunction has shown to be levodopa-responsive in PD in a subset of patients (19).

When analyzing speech parameters in Parkinsonian syndromes, it is therefore of essential importance to take motor impairment into account, which has not been included in previous studies (8–14).

Moreover, it is recommended to use multiple speech tasks to assess dysarthria in movement disorders (20), which leads to an abundance of potential parameters with time-consuming recordings and evaluation procedures. So far it remains unclear which speech characteristics are valid digital biomarkers in delineating MSA from PD.

In this work, we present results from a study conducted at two German centers analyzing speech recordings of patients with MSA and PD, and investigating the factors of speech that characterize both diseases. In addition, we analyzed the relationship between motor impairment measured by the Movement Disorders Society-Unified Parkinsons’s disease rating scale (MDS-UPDRS) part III (21) and speech characteristics for MSA and PD. Based on these results we developed statistical models for delineating MSA from PD based on speech characteristics.

## Methods

### Recruitment of study participants

The study was approved by the institutional review board of Technische Universität Dresden, Germany (BO-EK-149032021, BO-EK-47012020) and the Brandenburg State Medical Association (S21(a)/2017). Between March 2021 and November 2022, 23 people with PD according to the MDS diagnostic criteria for PD (22) and 21 people with probable or possible MSA according to the MDS diagnostic criteria for MSA (23) were recruited at two German movement disorder centers. Written informed consent was obtained from all participants. MDS-UPDRS III was obtained from all patients (21). Unified Multiple System Atrophy Rating Scale (UMSARS) was obtained from all MSA patients (24).

### Speech recordings

Recordings were performed in a quiet room with low ambient noise using an Olympus Linear PCM Recorder LS-P4 and a mouth-to-microphone distance of 5 cm. The recordings were processed with 16 bit resolution and 44.1 kHz sample size. Artificial recording alterations such as equalization, automatic gain control, and noise reduction were disabled.

### Speech tasks

Recordings were made for the following tasks: (1) sustained vowel phonation of the vowels /a/ and /i/, (2) sequential motion rates /p?t?k?/, (3) alternating motion rates /p?/ and /t?/, and (4) text reading. Recordings for each task were performed twice and closely followed the *Guidelines for Speech Recording and Acoustic Analysis in Dysarthrias of Movement Disorders* (20).

### Acoustic analysis

Based on the *Guidelines for Speech Recording and Acoustic Analysis in Dysarthrias of Movement Disorders (20)*, we calculated a set of speech characteristics (Table S1) using Praat (25): (1) *total pause duration*, (2) *mean pause duration*, (3) *intensity variability*, (4) *F0*, (5) *F0 variability*, (6) *harmonics-to-noise ratio (HNR)*, (7) *jitter*, (8) *shimmer, (9) vowel space area (VSA)* and (10) *voice breaks*.

The speech characteristics (1) *reading duration*, (2) *maximum phonation time*, (3) *syllable duration*, (4) *syllable count*, (5) *rhythm acceleration*, (6) *rhythm instability*, (7) *voice onset time (VOT)* and (8) *VOT variability* were calculated based on manual inspection and annotation of the sound files in Praat. For robust measurements to outliers, we used the median syllable length, median VOT and median absolute deviation of VOTs. Rhythm acceleration and instability were calculated by modeling a linear increase or decrease in speech rhythm (10). More details are provided in the supplement.

To increase the robustness of our measurements and to limit the number of speech characteristics to a reasonable number, the following steps were taken: All speech tasks and calculations described above were performed twice, and only the means of both recordings were analyzed. For sustained phonation, the means of the speech characteristics from the /a/ and /i/ tasks were calculated. Also, means of the speech characteristics of /p?/, /t?/ and /p?t?k?/ were calculated.

To account for potential sex differences in speech, the following steps were taken: *F0* was analyzed separately for male and female patients due to the higher pitch in females. For all other speech characteristics, male and female patients were initially analyzed together. Speech characteristics that showed significant differences between males and females in MSA or PD were subsequently analyzed in sex-specific subgroups.

### Statistical analyses

Exploratory factor analysis using a minimum residual solution and no factor rotation was performed on all MSA and PD patients with complete recordings, based on all speech characteristics except *F0*, as this parameter is highly influenced by sex. To account for the smaller number of MSA patients who completed all tasks, we selected an age- and sex-matched subgroup of the PD cohort of equal size for the exploratory factor analysis. The number of three factors was determined visually from the scree plot (Fig. S1).

MSA and PD speech characteristics, MDS-UPDRS III, age, and disease duration were compared using t-test, Welch’s test or Mann-Whitney-U test, depending on the distribution of the data. Sex was compared using Fishers exact test and Hoehn & Yahr (H&Y) using Kolmogorov-Smirnov test. Additionally, MSA and PD speech characteristics were compared using a linear model with MDS-UPDRS III as covariate. Observations with an absolute z-score greater than 3 were removed for statistical tests to limit the effect of outliers.

To explore the potential for discriminating MSA from PD, logistic regression models using different predictors were analyzed: (I) a null model including only MDS-UPDRS III, (II) models including only a single speech characteristic or speech factor, and (III) models including MDS-UPDRS III and either one speech characteristic or one speech factor. Only speech characteristic with significant differences between MSA and PD after correcting for MDS-UPDRS III were considered.

All tests were conducted as two-tailed with a significance level 0.05. Statistical analysis were performed using Python 3.10.8 (26) with the packages scipy 1.9.3 (27), factor-analyzer 0.4.1 (28), pingouin 0.5.2 (29), statsmodels 0.13.2 (30), and scikit-learn 1.0.2 (31).

### Code availability

The complete Praat scripts used for our analyses are available at github.com/t-haehnel/MSA-Speech-Analysis-Praat.

## Results

### Characteristics of study participants with MSA and PD

Speech recordings from 21 MSA patients and 23 PD patients were included in this study. Text reading recordings were available for all patients. Diadochokinetic tasks were recorded in 13 MSA patients and 22 PD patients. Sustained phonation was recorded in all PD patients and 13 MSA patients. Based on these recordings, we calculated the voice characteristics described in Table S1.

The clinical characteristics of the MSA and PD cohorts are described in Table 1. As expected, MSA patients had a shorter disease duration (p=0.001), but higher motor impairment reported by MDS-UPDRS III (p=0.001) and disease severity on the Hoehn & Yahr scale (p=0.001).

**Table 1:**
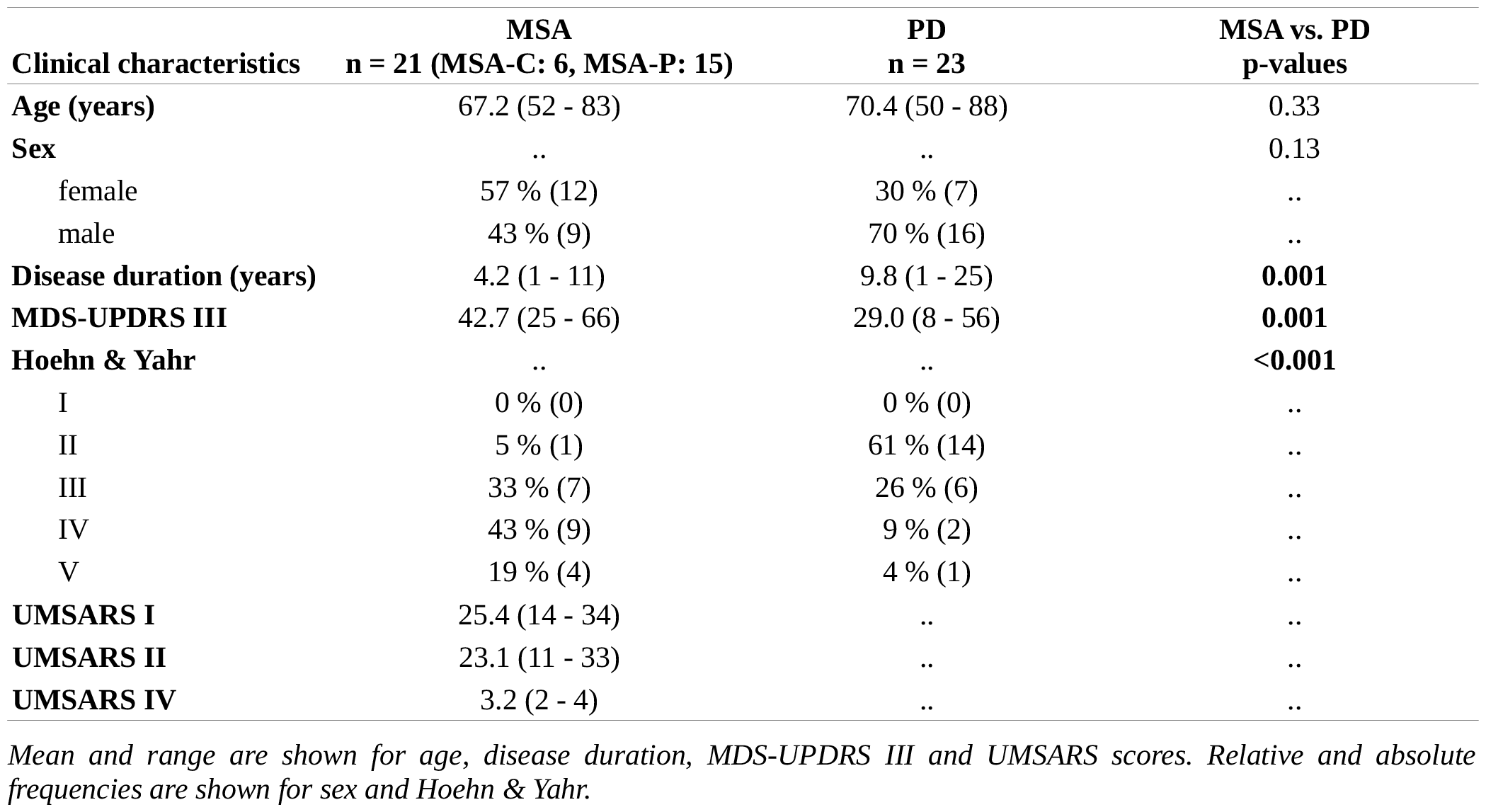
Clinical characteristics of the MSA and PD cohorts.

Sex-specific *F0* differences were observed, as expected, for the reading task (PD: p<0.001; MSA: p<0.001) and the sustained phonation tasks (PD: p<0.001; MSA: p=0.001). No sex-specific differences were found for other speech characteristics in MSA patients. For patients with PD, most speech characteristics were not affected by sex, except for a higher *number of pauses* (p<0.001), a shorter *mean pause duration* (p<0.001), and a higher *vowel space area* (p=0.02) in female PD patients compared to male PD patients.

### Speech factors

We performed an exploratory factor analysis to identify common factors underlying the speech recordings (Fig. 1). As recommended, only factor loadings with an absolute value greater than 0.63 were considered relevant (32).

**Figure 1:**
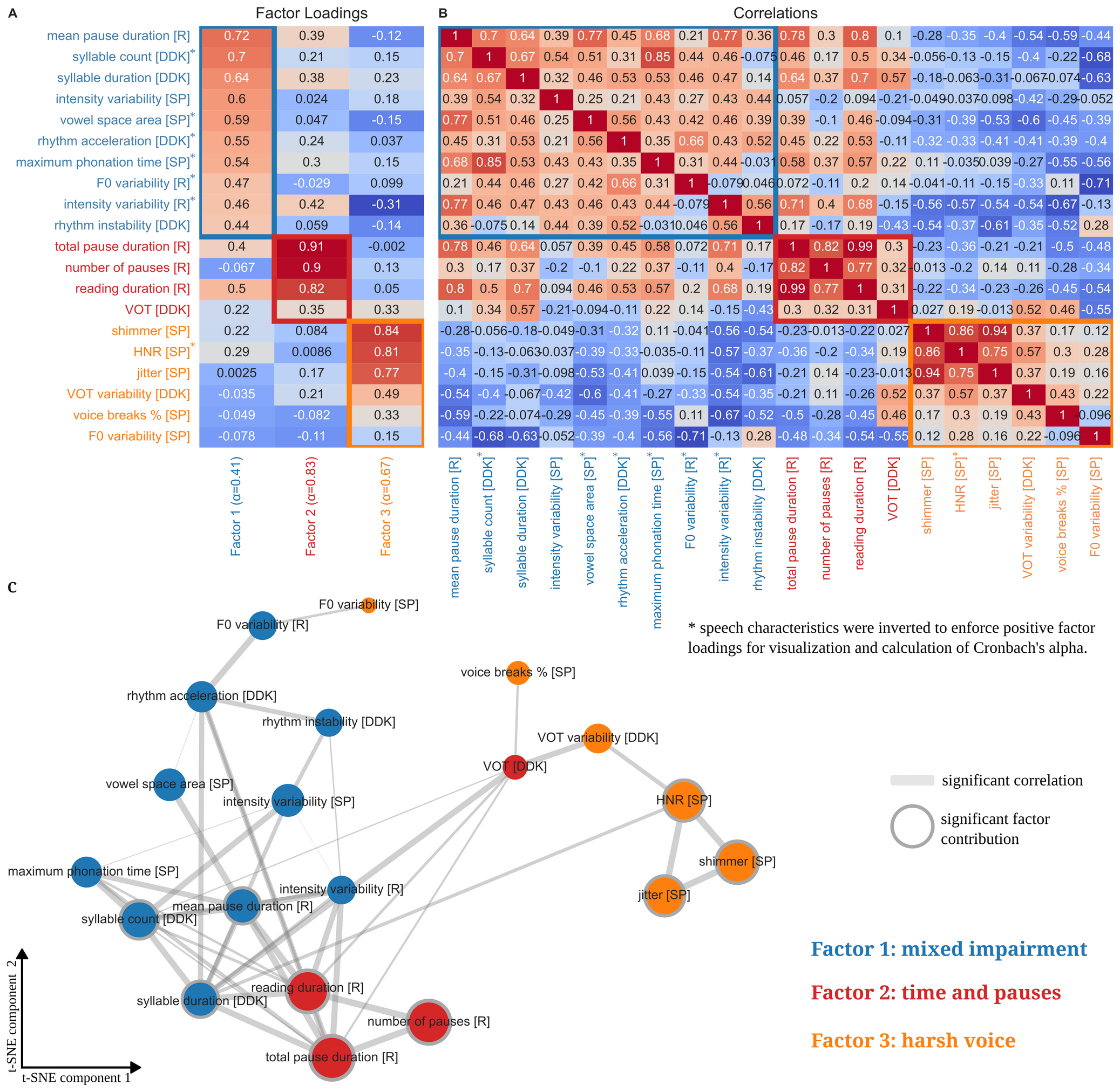
MSA and PD speech factors and correlations. Factor loadings (A) and correlations (B) of speech characteristics. C: t-SNE with significant correlations as gray lines (thickness corresponding to correlation coefficients) and relevant factor contributions as gray surroundings.

The first factor consisted of 10 different speech characteristics obtained from all three speech recording tasks. Cronbach’s alpha showed an unacceptable internal consistency (α=0.41), and most of the speech characteristics had low factor loadings below 0.63. Thus, only the following factors with loadings above 0.63 were considered relevant: *mean pause duration, syllable count*, and *syllable duration*. Due to the high number of different speech characteristics and the low internal consistency, we summarized this factor as *mixed speech characteristics*. The second factor presented good internal consistency (α=0.83) and consisted of four speech characteristics, three of them considered relevant: *total pause duration, number of pauses, reading duration*. Therefore, we refer to the second factor as *time and pauses*. The third factor consisted of six speech characteristics of which three were considered relevant: *shimmer, HNR* and *jitter*. All of these characteristics are obtained from the sustained phonation speech task and are markers of *harsh voice*. Also, two of the speech characteristics that were not considered relevant are markers of harsh voice and are obtained from the sustained phonation task: *F0 variability* and *voice breaks*. The internal consistency of this factor was questionable (α=0.67).

A comparable clustering pattern between speech characteristics was observed based on t-distributed Stochastic Neighbor Embedding (t-SNE) (Fig. 1C).

### Speech characteristics as marker of motor impairment

We aimed to investigate the influence of motor impairment on speech characteristics in PD and MSA. Therefore, we correlated the speech characteristics listed in Table S1 and the speech factors identified above with motor impairment as reported by MDS-UPDRS III (Fig. 2, Table S2). For both MSA and PD, this revealed a significant and strong correlation with *F0 variability* of the sustained phonation task. In addition, we observed an association of impaired *rhythm instability* with more severe motor impairment in MSA patients which we did not observe in the PD cohort. In addition, several other speech characteristics were correlated with MDS-UPDRS III in the PD cohort (Fig. 2).

**Figure 2:**
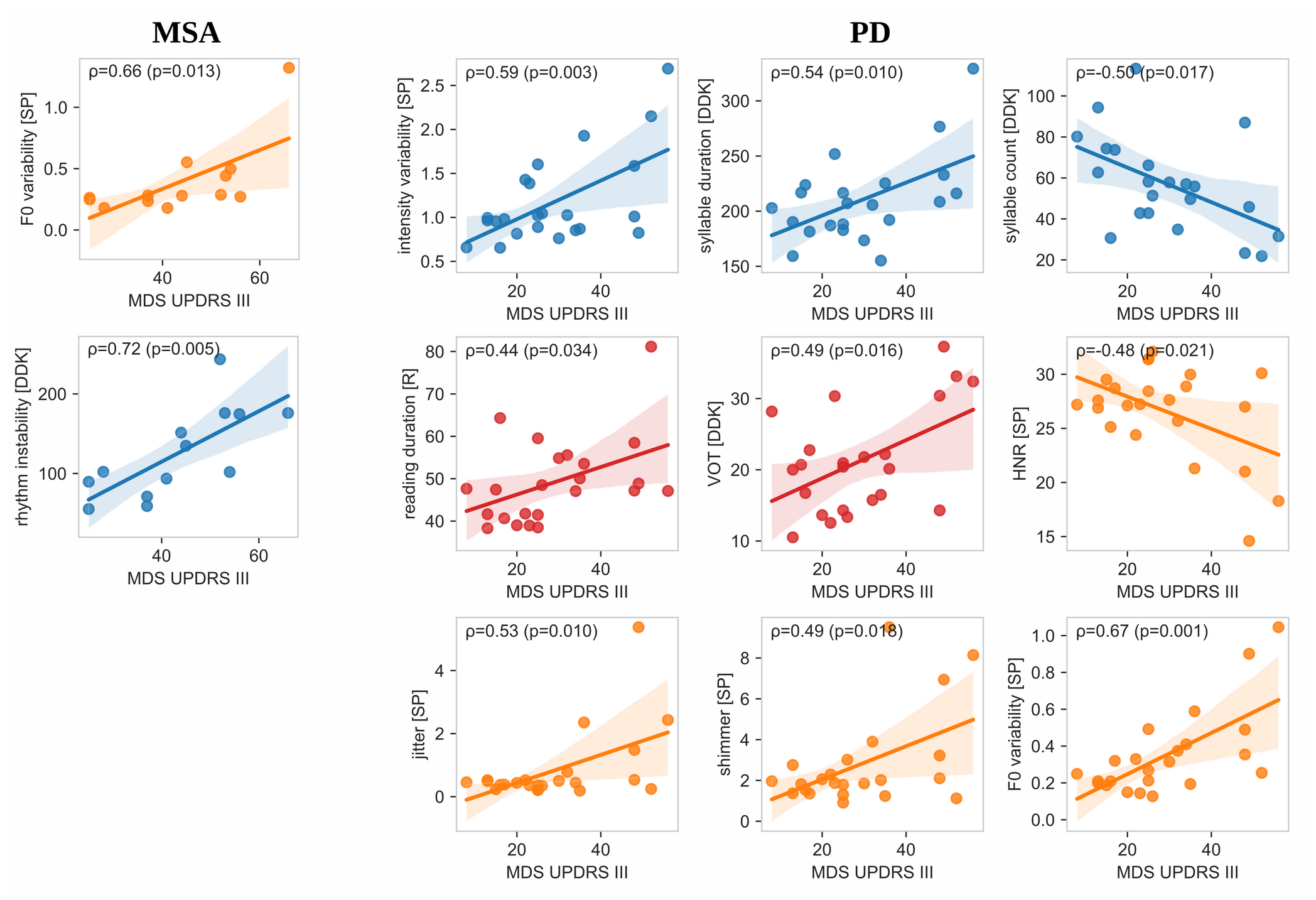
Speech characteristics as markers of motor impairment in MSA and PD. Significant correlations of MSA and PD speech characteristics with motor impairment reported by MDS-UPDRS III. Pearson correlation coefficients, corresponding p-values and 95% confidence intervals are shown.

### Differentiating MSA and PD using speech characteristics

When analyzing speech differences between MSA and PD without including MDS-UPDRS-III correction, we identified significant differences in several speech characteristics from all speech tasks and all speech factors (Fig. S2-S5 and Table S2). Specifically, we found longer *VOTs* (p=0.01) with higher *VOT variability* (p=0.009), higher *rhythm instability* (p=0.004) with lower *rhythm acceleration* (p=0.049) and a longer *syllable duration* (p=0.02) for the diadochokinetic tasks in the MSA cohort compared to PD. When analyzing the sustained phonation task, we observed higher *intensity variability* (p=0.04) in the MSA cohort. Analysis of the reading task revealed lower *number of pauses* (p=0.007), lower *intensity variability* (p=0.04) and lower *F0 variability* (p=0.03) in the MSA cohort.

Sex-specific subgroup analyses were performed for the three speech characteristics with significant differences between male and female MSA patients. Thereby, we confirmed the *lower number of pauses in* both female (p=0.001) and male (p=0.02) MSA patients compared to PD patients. In contrast, *mean pause duration* showed no differences between MSA and PD in the overall analysis and in male patients, but female MSA patients presented higher values (p=0.01). No differences were observed in the sex-specific subanalyses of *vowel space area*.

Comparing *F0* values for both reading and sustained phonation task, we found higher F0 values for both male and female MSA patients, although the differences were not significant (Fig. S6).

Regarding speech factors, we observed higher *mixed impairment* (p=0.016) and higher *harsh voice* (0.04) in the MSA cohort, whereas *time and pauses* were more impaired (p=0.006) in the PD cohort.

To correct for the significant contributions of motor impairment to speech characteristics, as shown in Fig. 2, we next implemented the MDS-UPDRS III as a correction factor. Using this model, we identified four voice characteristics and one speech factor that were significantly different between MSA and PD (Fig. 3, Table S2). After correction for motor impairment, MSA patients exhibited a higher *rhythm instability* (p=0.002), longer *syllable duration* (p=0.03) and lower *number of pauses* (p=0.003) compared to PD. In addition, MSA patients presented a lower corrected *F0 variability* for the sustained phonation task (p=0.01) which was not visible in the univariate analysis before. Regarding speech factors, we observed a significant difference with less *time and pauses* impairment (p=0.006) in the MSA cohort.

**Figure 3:**
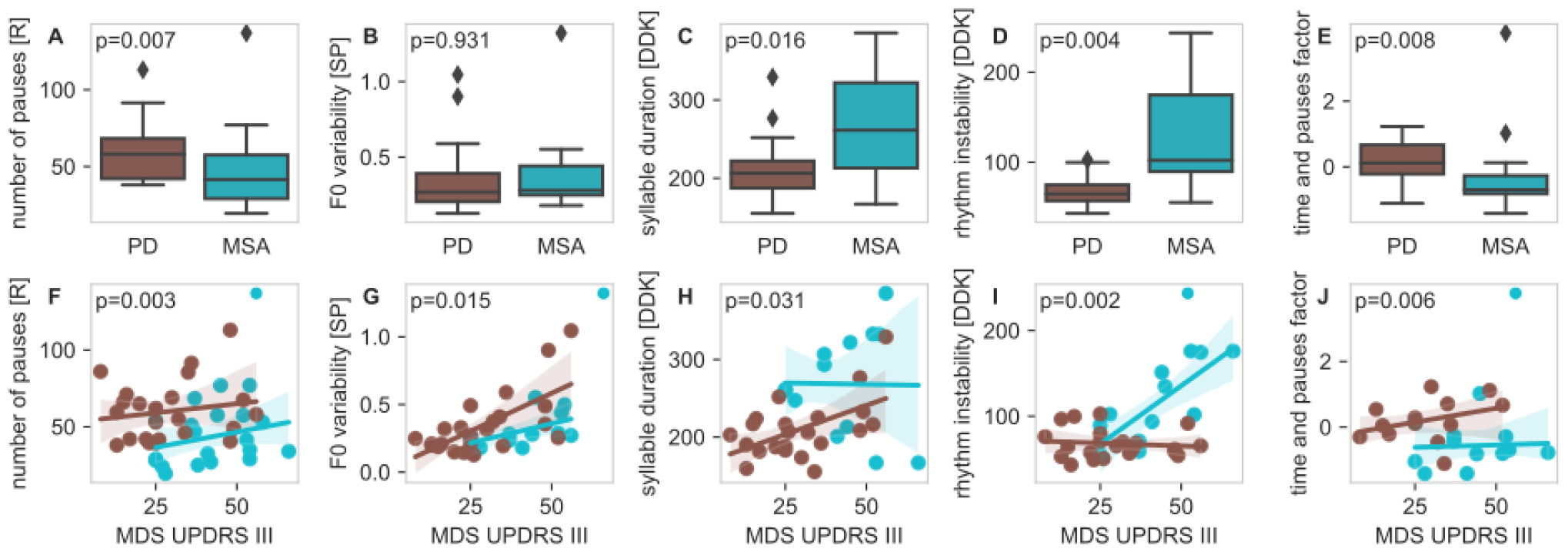
Speech differences between MSA and PD. All speech characteristics and speech factors with significant differences between MSA (blue) and PD (brown) in the MDS-UPDRS III corrected analysis. Univariate comparison (top) and comparisons corrected by MDS-UPDRS III (bottom) are shown.

Again, sex-specific subgroup analyses were performed for the three speech characteristics with significant differences between male and female MSA patients. The lower *number of pauses* was also significant in the female (p<0.001) and male (p=0.01) MSA subgroups. Consistent with the overall analysis, *vowel space area* showed no sex-specific differences between the groups. In addition, we observed a significantly higher *mean pause duration* in male (p=0.05), but not female MSA patients.

Finally, we investigated the potential for discriminating between MSA and PD based on the four speech characteristics and one speech factor that showed significant differences between MSA and PD. The best discrimination was observed for *syllable duration* with an AUC-ROC of 0.89 when MDS-UPDRS III was also included. In general, all models improved after inclusion of MDS-UPDRS III (Fig. 4).

**Figure 4:**
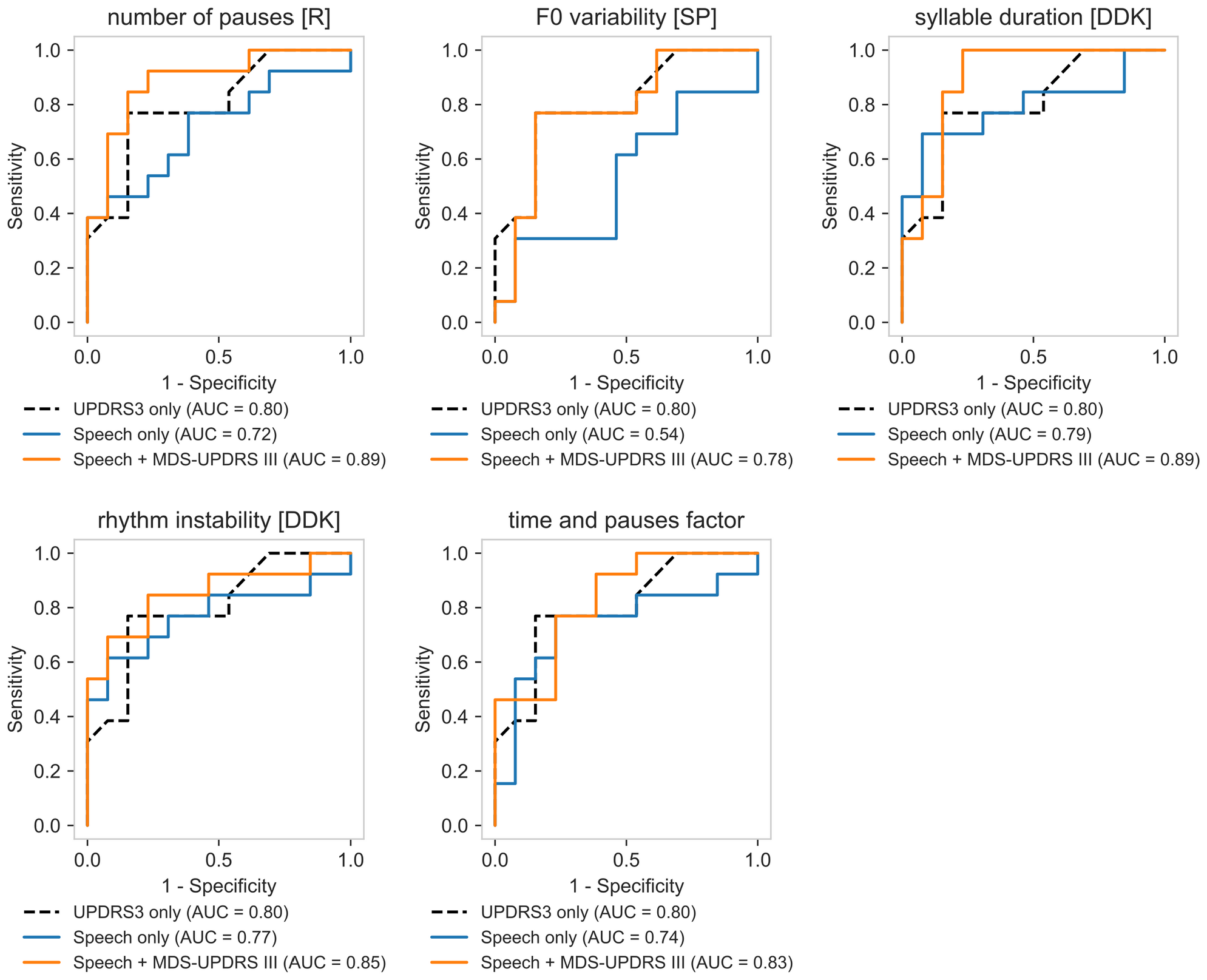
Differential diagnosis between MSA and PD. Comparison of receiver operator characteristic (ROC) curves for different logistic regression models.

## Discussion

### Principal Results

This is the first study on speech analysis in German speaking MSA patients, while speech characteristics have been studied in Czech (8,10,11,14) and Italian (13) speaking MSA patients.

The speech differences we observed between MSA and PD, when not correcting for motor impairment, are in line with previous results (8–11,13,14), proving the validity of our analysis and demonstrating that our findings are valid across different languages. More specifically, similar findings have been reported by others for *F0 variability* in the reading task (13), *intensity variability* in the sustained phonation task (9), *syllable duration* (13,14), *rhythm instability* (9,10,13), *VOT* (13) and *VOT variability* (14), showing more severe speech impairment in the MSA cohort when compared to PD. Furthermore, our results are consistent with previous findings that male MSA patients have higher F0 than male PD patients (12) and suggest that F0 may also be higher in female MSA patients compared to female PD patients. We observed less *F0 variability* in the sustained phonation task in MSA when correcting for MDS-UPDRS III, whereas several studies reported higher impairment in MSA compared to PD in an MDS-UPDRS III uncorrected analysis (9,11,13). Furthermore, we found a higher *rhythm acceleration* and higher *number of pauses* in PD patients compared to MSA, which has not been reported before.

Based on speech recordings of native German speaking MSA and PD patients, we identified two underlying speech factors with acceptable internal consistency and high clinical interpretability, as well as a third factor of mixed speech characteristics. We were able to discriminate MSA and PD using a single speech characteristic, while optimal results were obtained when MDS-UPDRS III scores were also included in the analysis. We provide our speech analysis scripts along with this publication for other researchers to facilitate more standardized speech analysis.

### Speech Factors

We identified two underlying speech factors with high internal consistency that can be interpreted as distinct speech domains: *time and pause* characteristics and *harsh voice*. These domains were also visually confirmed using t-SNE as an alternative approach. Interestingly, the *time and pause* factor consisted mainly of parameters from the reading task, whereas the *harsh voice* factor consisted mainly of parameters from sustained phonation.

Previous publications on MSA speech distinguished three domains of speech impairment based on neuropathological constructs and perception: hypokinetic, ataxic and spastic (8,11,13,15). In general, our study did not confirm this “syndromal” classification from a data-driven perspective. Hypokinetic, ataxic, and spastic characteristics were distributed along all three speech factors and did not show an association with any of them. Furthermore, most of the speech characteristics that are traditionally interpreted as hypokinetic characteristics showed no correlation with MDS-UPDRS III in PD patients (i.e., *number of pauses, mean pause duration, F0 variability of reading task, maximum phonation time, vowel space area, rhythm acceleration*). In MSA, none of the traditional hypokinetic speech characteristic correlated with MDS-UPDRS III. In contrast, two traditional ataxic speech characteristics (*F0 variability* of sustained phonation and *rhythm instability*) correlated with MDS-UPDRS III.

Our results show that there is no single speech task that can capture all three speech factors. Thus, it remains essential to perform different tasks to capture the full set of speech characteristics in MSA and PD.

### Speech characteristics as marker of motor impairment

While speech in MSA is known to also exhibit hypokinetic pathology (8,10,13,15), it has not been investigated whether speech characteristics can directly report the severity of motor impairment. Our work has shown that *F0 variability* in sustained phonation and *rhythm instability* are both strongly correlated with motor impairment in MSA. An association of higher *F0 variability* with more severe motor impairment has also been observed in PD patients from our cohort and by others (33). In contrast, most speech characteristics were not associated with motor impairment in MSA, demonstrating that speech characteristics contain additional clinical information that cannot be captured by this clinical score, highlighting the importance of acoustic speech analysis in movement disorders.

Speech characteristics have been extensively studied as a marker of disease progression in PD (34–37). Longitudinal studies have shown that markers of rhythm instability and rhythm acceleration (35), *vowel space area* and *vowel articulation index* (36), *shimmer, HNR*, and several pause characteristics show significant changes with PD progression (37). In contrast, whether speech characteristics are directly correlated with motor impairment as reported by UPDRS III remains controversial. While some studies reported significant correlations of prosodic parameters (33,38), several studies found no relationship with motor impairment (33,35–37,39,40). Our results support previous findings (33,38) that *jitter, shimmer, HNR*, and *F0 variability* of the sustained phonation task report motor impairment, as we also observed correlations with MDS-UPDRS III for these speech characteristics. Beyond these known associations, we show that *intensity variability* of the sustained phonation task, *syllable duration, syllable count, reading duration and VOT* also capture motor impairment.

### Speech differences in MSA and PD: correcting for motor impairment

As expected (16), the severity of motor impairment was higher in our MSA cohort compared to our PD cohort. This led us to question whether the speech differences observed by us and others are disease specific or at least partly due to differences in motor impairment.

Previous studies did not consider the more pronounced motor impairment in MSA compared to PD (9– 11,13,14), thus potentially introducing an important bias into their analyses. In fact, our results show that several speech differences between MSA and PD can be explained by the more severe motor impairment in MSA compared to PD and disappear after correction for MDS-UPDRS III. Therefore, the four speech characteristics that remain significant after correction for MDS-UPDRS III are more likely to capture true disease-specific speech characteristics and should be used to differentiate MSA and PD. Specifically, we observed lower *number of pauses*, lower *F0 variability* in sustained phonation, higher *syllable duration*, and higher *rhythm instability* in MSA patients after correcting for motor impairment.

Furthermore, the importance of taking motor impairment into account when discriminating MSA from PD based on speech features is also supported by the higher AUC-ROC scores when including MDS-UPDRS III in our predictive models.

We demonstrated that MSA can be differentiated from PD with good accuracy using only MDS-UPDRS III and one speech parameter as predictors, thereby achieving a comparable performance as reported by others (13). In general, this outlines the value of speech assessments for differential diagnosis in movement disorders.

### Limitations

The number of MSA patients in our analysis is comparable with most acoustic studies in MSA, which included 9 to 30 MSA patients (8–12) and below one study including 40 MSA patients (13). Still, the number of individuals is limited, and it is likely that small differences in acoustic parameters between MSA and PD were not recognized. Furthermore, we did not perform an MSA subtype analysis due to the small number of cerebellar MSA patients in our cohort (n=6). Recently published work suggests that at least some speech characteristics differ between MSA subtypes which needs to be validated in other languages and larger cohorts (13).

Another limitation is the small number of MSA patients with low MDS-UPDRS III (at least 25 points in our cohort). Although this is expected due to the faster disease progression in MSA and the difficulty of differential diagnosis in an early disease stages (18), our results currently prevent us from delineating both diseases in a very early disease stages. The revised MDS diagnostic criteria for MSA have now implemented research criteria for possible prodromal MSA (41), allowing future studies to focus on speech analysis in a very early disease stage.

## Conclusion

Our study increases the evidence that speech assessments provide important biomarkers for differential diagnosis between MSA and PD. The identification of meaningful factors that characterize speech in patients with PD and MSA will allow comparison of findings across different studies and allow researchers to simplify speech assessments in subsequent studies, thereby reducing participant burden. We emphasize the importance of correcting for motor impairment when analyzing speech characteristics to avoid an important bias in speech analysis for differential diagnosis of Parkinson syndromes. We provide a set of four speech parameters that should be used to discriminate MSA from PD. Furthermore, our results challenge the traditional classification of hypokinetic, ataxic, and spastic speech factors. We show that different speech tasks are required to capture all underlying factors of dysarthria in MSA, which are not well reflected by commonly used clinical scores like MDS-UPDRS III. Further research should especially focus on early MSA disease stages and MSA subtypes.

## Data Availability

All Praat scripts used for our analyses are available at github.com/t-haehnel/MSA-Speech-Analysis-Praat. Speech recordings can not be made available to others due to privacy reasons.

## Acknowledgment

We thank all patients for their participation in the study.

## Author’s roles

Design: TH, BF, FG; Execution: TH, AN, KS, LB, AV; Analysis: TH; Writing: TH, BF, FG.

