## Supplemental Material for "Speech Differences between Multiple System Atrophy and Parkinson’s Disease: a Multicenter Study"

### Additional figures and tables

| Speech characteristic | Speech task | Unit | Definition |
|---|---|---|---|
| Reading duration | Reading | s | Total duration required for text reading passage including pauses |
| Total pause duration | Reading | s | Sum of all pause segments with a duration longer than 60 ms |
| Number of pauses | Reading | - | Number of pauses with a duration longer than 60 ms (1) |
| Mean pause duration | Reading | ms | Mean duration of pauses |
| Intensity variability | Reading | dB | Standard deviation of loudness |
| F0 | Reading | Hz | Fundamental frequency (F0) |
| F0 variability | Reading | semitones | Standard deviation of fundamental frequency (F0) |
| Harmonics-to-noise ratio (HNR) | Sustained phonation | dB | Degree of acoustic periodicity in the voice signal |
| Jitter | Sustained phonation | % | Period-to-period variability in fundamental frequency (F0) |
| Shimmer | Sustained phonation | % | Period-to-period variability of amplitudes |
| Intensity variability | Sustained phonation | dB | Standard deviation of loudness |
| F0 | Sustained phonation | Hz | Fundamental frequency (F0) |
| F0 variability | Sustained phonation | semitones | Standard deviation of fundamental frequency (F0) |
| Maximum phonation time (MPT) | Sustained phonation | s | Total duration of phonation time |
| Vowel space area (VSA) | Sustained phonation | - | Vowel space area calculated as Euclidean distance between the first and second formant of the vowels /a/ and /i/ in the formant space (2) |
| Voice breaks | Sustained phonation | % | Proportion of subharmonic intervals |
| Syllable duration | Diadochokinetic | ms | Median syllable length |
| Syllable count | Diadochokinetic | - | Number of syllables per diadochokinetic task |
| Rhythm acceleration | Diadochokinetic | ‰ | Acceleration of syllable duration over the task (in ms acceleration per second speaking duration) (3) |
| Rhythm instability | Diadochokinetic | s | Sum of absolute deviations of each syllable duration from the expected duration divided by the total speech time. The expected syllable duration is calculated by taking the rhythm acceleration into account (3). |
| Voice onset time (VOT) | Diadochokinetic | ms | Duration from the release of the stop consonant until beginning of voicing |
| VOT variability | Diadochokinetic | ms | Median absolute deviation of the voice onset times |

***Table S1 List of speech characteristics calculated for the different speech tasks***

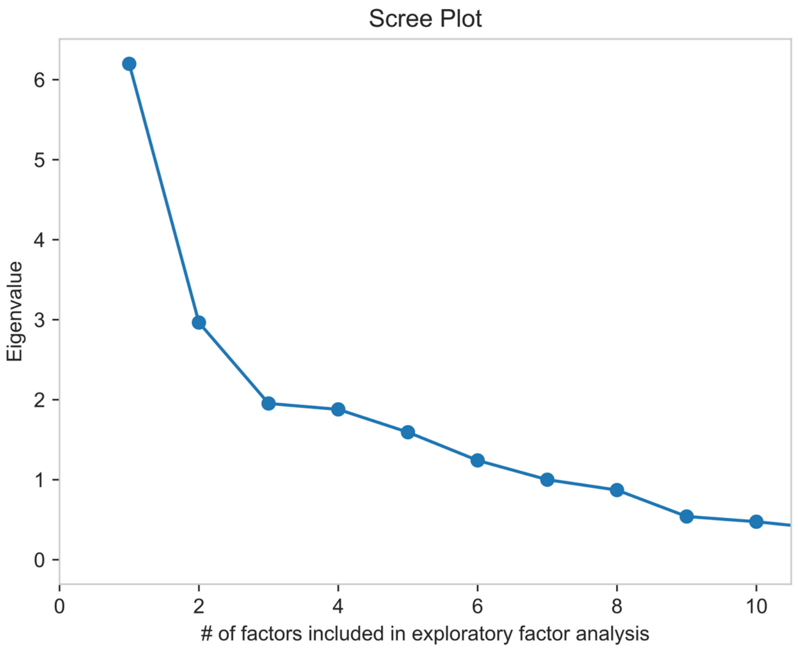

***Figure S1: Scree plot for exploratory factor analysis of speech characteristics***

*Scree plot showing the Eigenvalues for the exploratory factor analysis for 1 up to 10 factors. Based on this plot, k=3 factors were chosen as appropriate.*

| Speech characteristic or speech factor | Speech factor | MDS-UPDRS III correlation MSA cohort | MDS-UPDRS III correlation PD cohort | MSA/PD-Differences (not corrected) | MSA/PD-Differences (MDS-UPDRS III corrected) |
|---|---|---|---|---|---|
| **Mean pause duration [R]** MSA: male > female | **Mixed impairment** | p=0.87 | p=0.12 | p=0.12 | p=0.09 |
| female | | p=0.19 | p=0.08 | p=0.01 | p=0.40 |
| male | | p=0.45 | p=0.37 | p=0.20 | **MSA > PD (p=0.0497)** |
| **Syllable count [DDK]** | **Mixed impairment** | p=0.38 | **ρ=-0.50 (p=0.02)** | p=0.19 | p=0.72 |
| **Syllable duration [DDK]** | **Mixed impairment** | p=0.96 | **ρ=0.54 (p=0.010)** | **MSA > PD (p=0.02)** | **MSA > PD (p=0.03)** |
| **Intensity variability [SP]** | Mixed impairment | p=0.19 | **ρ=0.59 (p=0.003)** | **MSA > PD (p=0.04)** | p=0.93 |
| **Vowel space area [SP]** MSA: female > male | Mixed impairment | p=0.42 | p=0.28 | p=0.38 | p=0.57 |
| female | | p=0.58 | p=0.55 | p=0.30 | p=0.36 |
| male | | p=0.19 | p=0.11 | p=0.48 | p=0.62 |
| **Rhythm acceleration [DDK]** | Mixed impairment | p=0.24 | p=0.46 | **PD > MSA (p=0.049)** | p=0.10 |
| **Maximum phonation time [SP]** | Mixed impairment | p=0.42 | p=0.44 | p=0.39 | p=0.65 |
| **F0 variability [R]** | Mixed impairment | p=0.54 | p=0.47 | **PD > MSA (p=0.03)** | p=0.12 |
| **Intensity variability [R]** | Mixed impairment | p=0.18 | p=0.53 | **PD > MSA (p=0.04)** | p=0.20 |
| **Rhythm instability [DDK]** | Mixed impairment | **ρ=0.72 (p=0.005)** | p=0.63 | **MSA > PD (p=0.004)** | **MSA > PD (p=0.002)** |
| **Mixed impairment factor** | Mixed impairment | p=0.95 | p=0.055 | **MSA > PD (p=0.02)** | p=0.11 |
| **Total pause duration [R]** | **Time and pauses** | p=0.17 | p=0.05 | p=0.37 | p=0.10 |
| **Number of pauses [R]** MSA: female > male | **Time and pauses** | p=0.11 | p=0.45 | **PD > MSA (p=0.007)** | **PD > MSA (p=0.003)** |
| female | | p=0.20 | **ρ=0.78 (p=0.04)** | **PD>MSA (p=0.001)** | **PD > MSA (p<0.001)** |
| male | | p=0.35 | p=0.38 | **PD>MSA (p=0.02)** | **PD > MSA (p=0.01)** |
| **Reading duration [R]** | **Time and pauses** | p=0.16 | **ρ=0.44 (p=0.03)** | p=0.57 | p=0.90 |
| **VOT [DDK]** | Time and pauses | p=0.35 | **ρ=0.49 (p=0.02)** | **MSA > PD (p=0.01)** | p=0.08 |
| **Time and pauses factor** | Time and pauses | p=0.34 | p=0.25 | **PD > MSA (p=0.008)** | **PD > MSA (p=0.006)** |
| **Shimmer [SP]** | **Harsh voice** | p=0.16 | **ρ=0.49 (p=0.02)** | p=0.06 | p=0.73 |
| **HNR [SP]** | **Harsh voice** | p=0.85 | **ρ=-0.48 (p=0.02)** | p=0.05 | p=0.68 |
| **Jitter [SP]** | **Harsh voice** | p=0.41 | **ρ=0.53 (p=0.010)** | p=0.67 | p=0.09 |
| **VOT variability [DDK]** | Harsh voice | p=0.91 | p=0.05 | **MSA > PD (p=0.009)** | p=0.15 |
| **Voice breaks % [SP]** | Harsh voice | p=0.64 | p=0.08 | p=0.47 | p=0.14 |
| **F0 variability [SP]** | Harsh voice | **ρ=0.66 (p=0.01)** | **ρ=0.67 (p=0.001)** | p=0.93 | **PD > MSA (p=0.01)** |
| **Harsh voice factor** | Harsh voice | p=0.23 | p=0.95 | **MSA > PD (p=0.04)** | p=0.11 |

***Table S2: Summary of speech characteristics findings.***

*Summary of speech factor compositions, MDS-UPDRS III correlations, and differences between PD and MSA for all speech characteristics. Significance of differences between PD and MSA are shown with and without MDS-UPDRS III correction. Corresponding speech factors are shown in bold if the speech characteristic was considered relevant for the speech factor (i.e., factor loading greater than 0.63). Significant correlation coefficients and differences between MSA and PD are shown in bold. Pearson correlation coefficients are reported for significant correlations. Results of sex-specific subgroup analyses are shown for speech characteristics with significant differences between male and female patients.*

*Abbreviations: DDK: diadochokinetic task, HNR: harmonics-to-noise ratio, R: reading task, SP: sustained phonation task, VOT: voice onset time.*

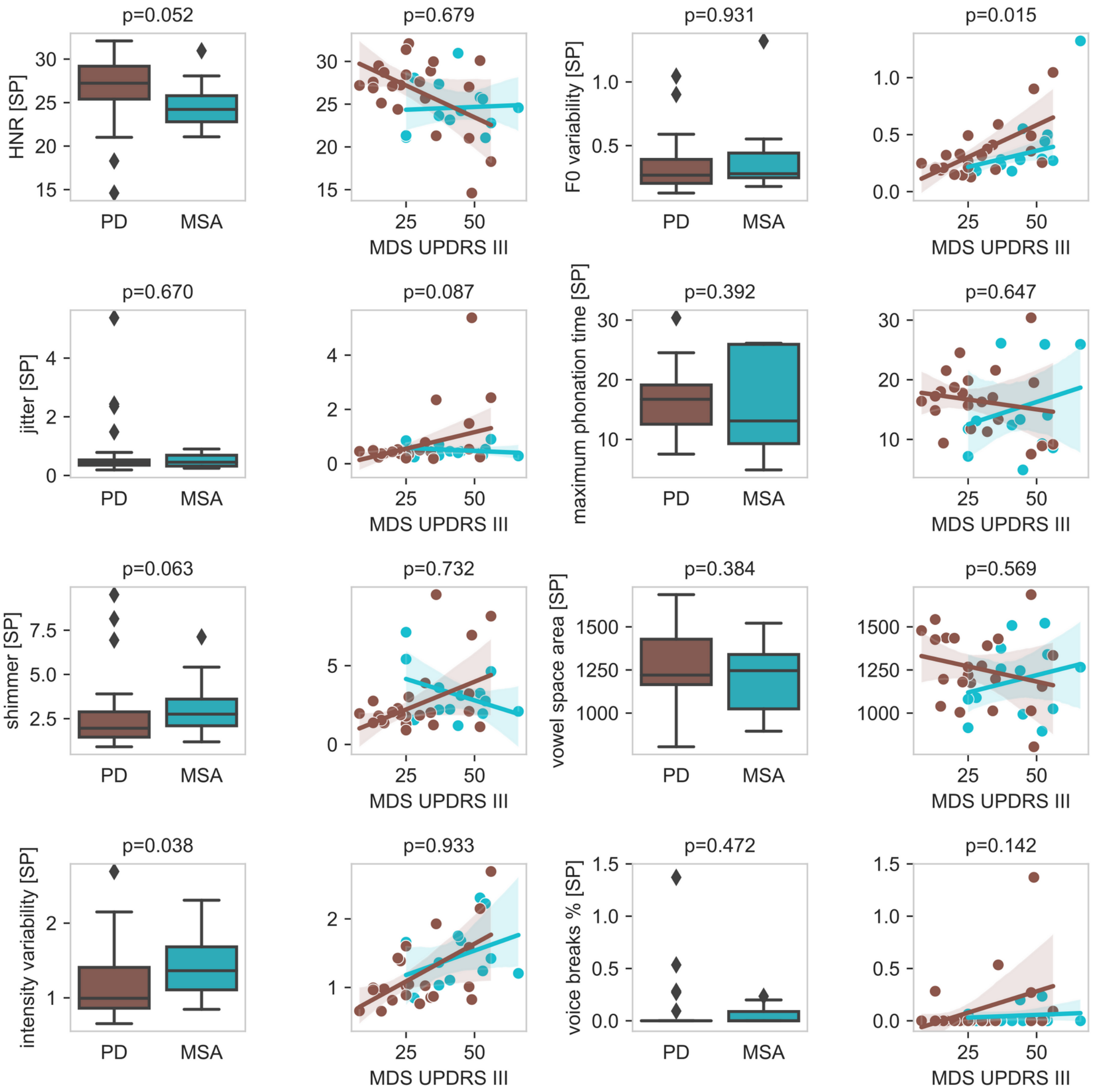

***Figure S2: Speech characteristics of MSA and PD for sustained phonation tasks***
*The left columns show the results of univariate comparisons between speech characteristics in PD and MSA with corresponding p-values. On the right columns, the parkinsonism severity reported by MDS-UPDRS III is included in the corresponding visualization. P-values for differential diagnosis reported on the right side were corrected by MDS-UPDRS III.*
*Abbreviations: HNR: harmonics-to-noise ratio, SP: sustained phonation task.*

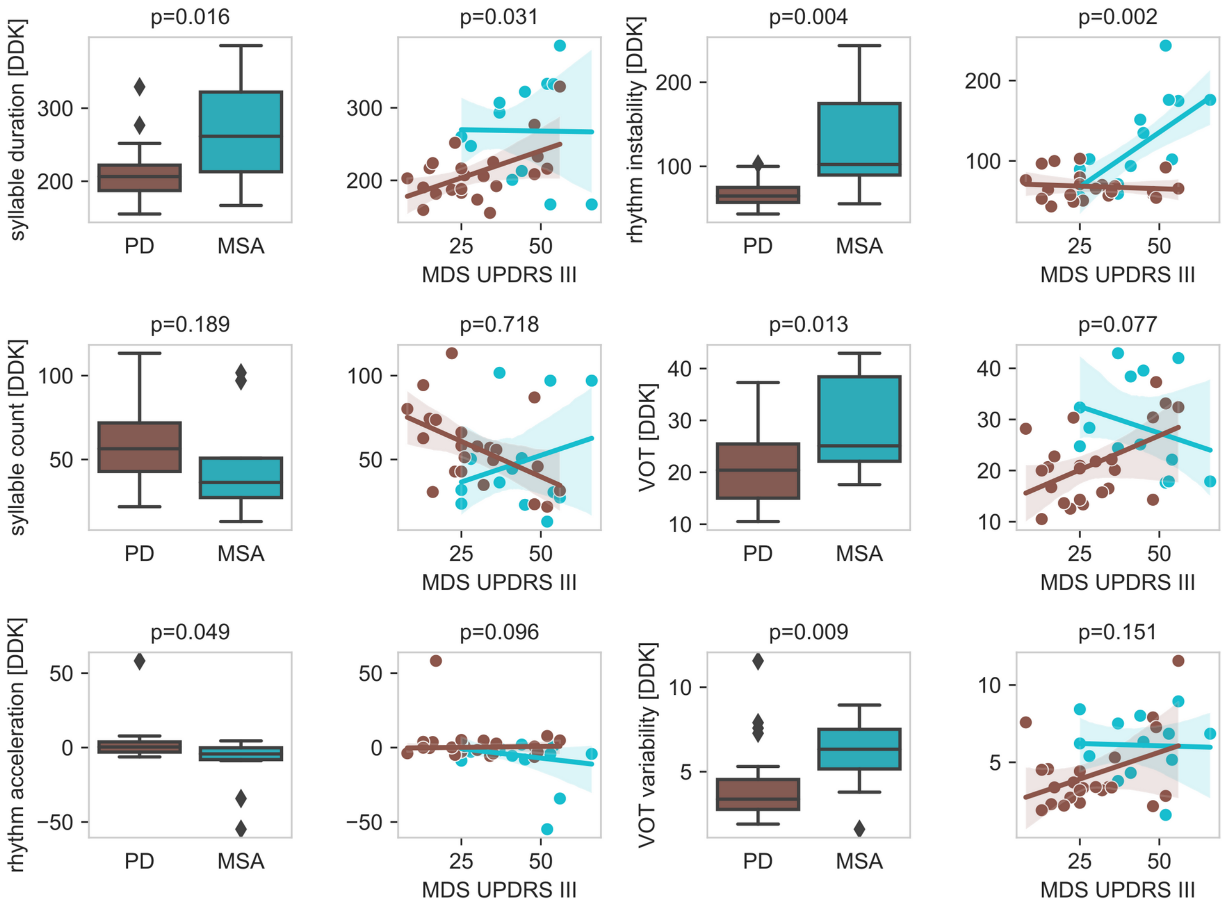

***Figure S3**: **Speech characteristics of MSA and PD for diadochokinetic tasks***
*The left columns show the results of univariate comparisons between speech characteristics in PD and MSA with corresponding p-values. On the right columns, the parkinsonism severity reported by MDS-UPDRS III is included in the corresponding visualization. P-values for differential diagnosis reported on the right side were corrected by MDS-UPDRS III.*
*Abbreviations: DDK: diadochokinetic task, VOT: voice onset time.*

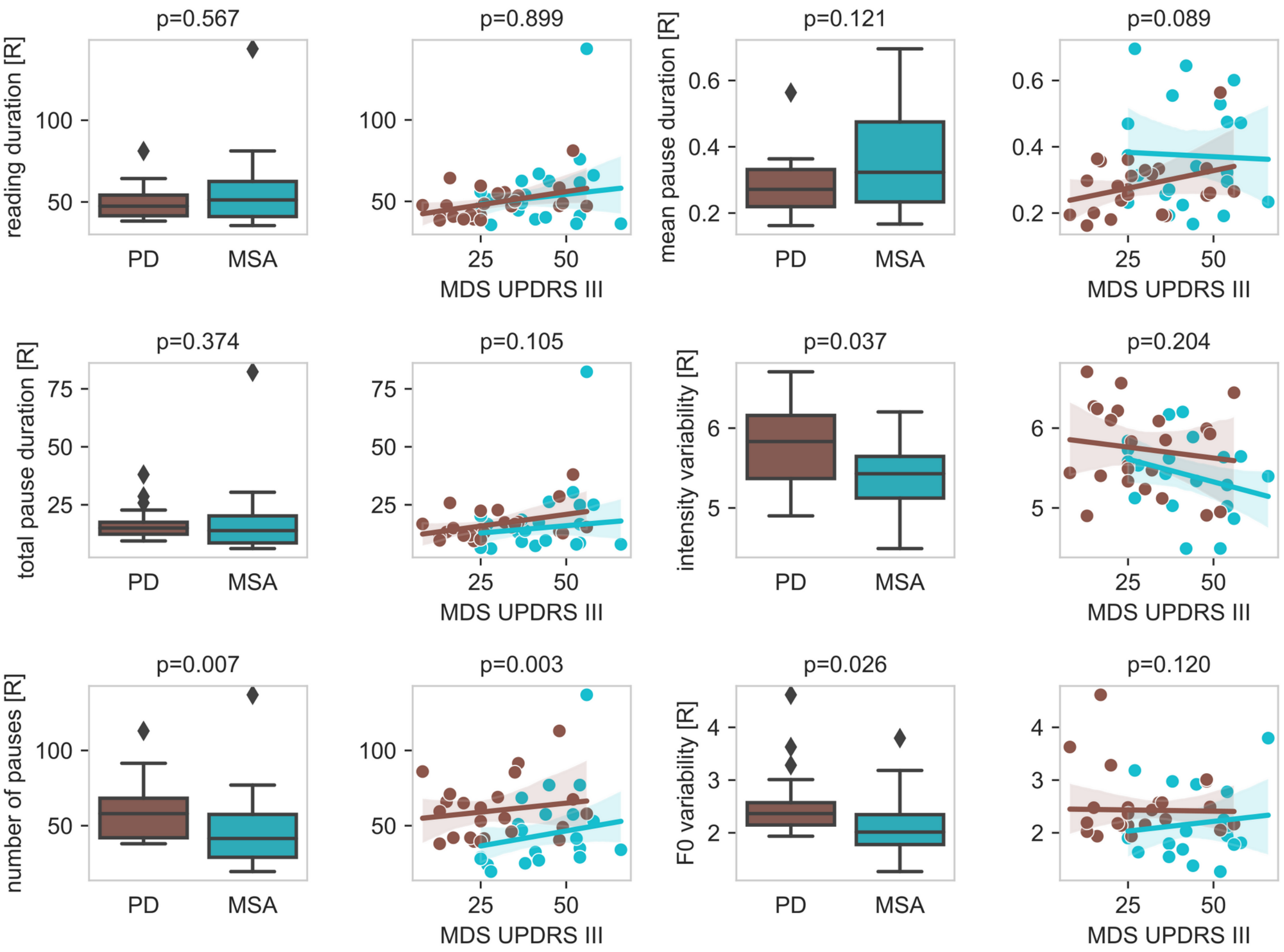

***Figure S4: Speech characteristics of MSA and PD for reading task***
*The left columns show the results of univariate comparisons between speech characteristics in PD and MSA with corresponding p-values. On the right columns, the parkinsonism severity reported by MDS-UPDRS III is included in the corresponding visualization. P-values for differential diagnosis reported on the right side were corrected by MDS-UPDRS III.*
*Abbreviations: R: Reading task*

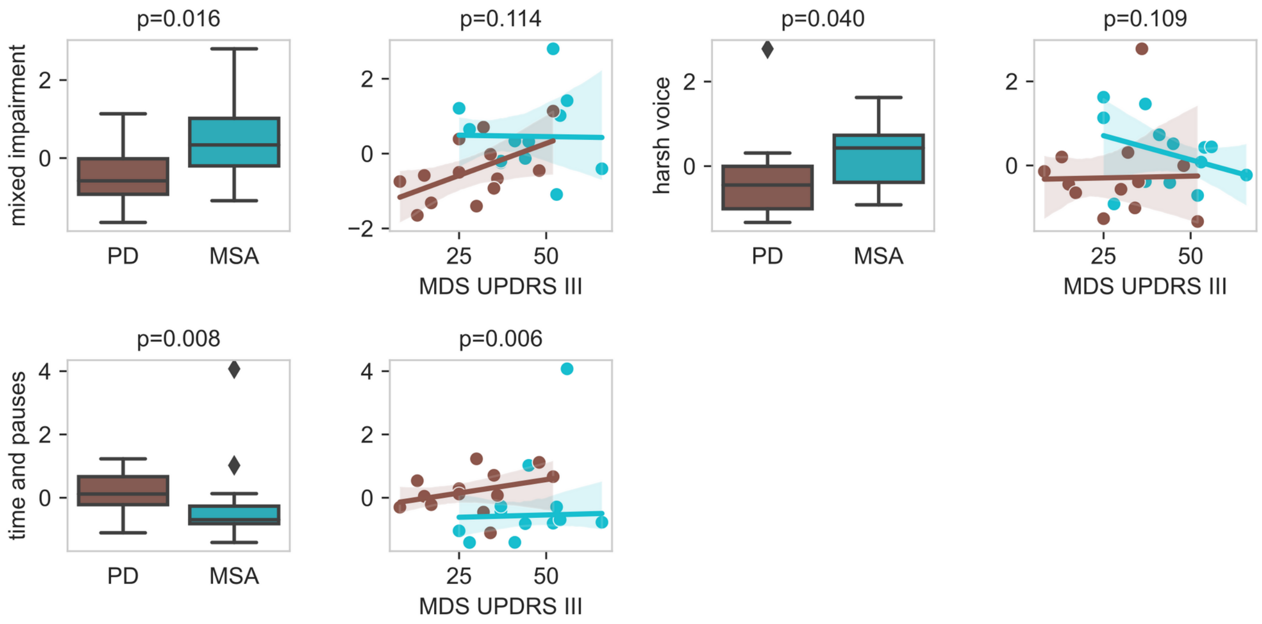

***Figure S5: Speech characteristics of MSA and PD for factors of the exploratory factor analysis***
*The left columns show the results of univariate comparisons between the speech factors in PD and MSA with corresponding p-values. On the right columns, the parkinsonism severity reported by MDS-UPDRS III is included in the corresponding visualization. P-values for differential diagnosis reported on the right side were corrected by MDS-UPDRS III.*

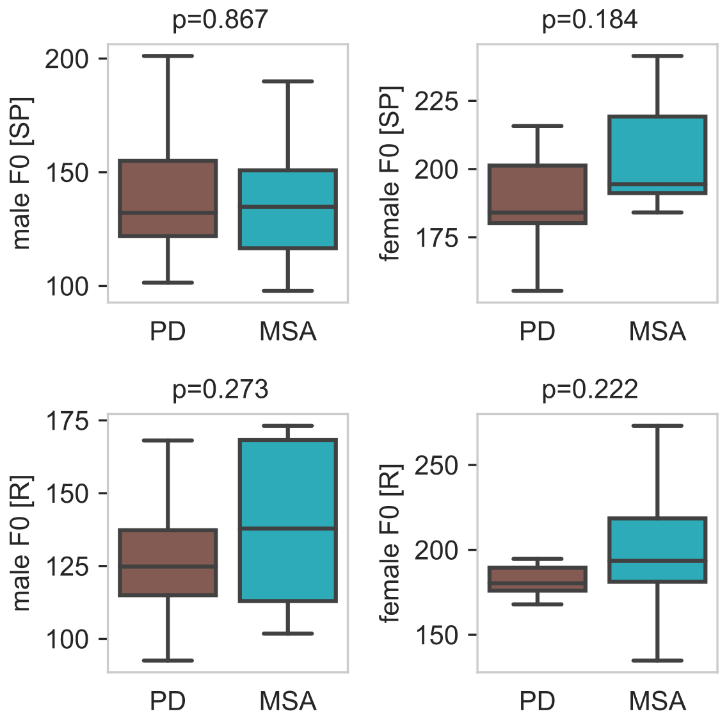

***Figure S6: Sex-specific analyses of F0 in MSA and PD***

*The figure shows the results of univariate sex-specific comparisons of F0 between PD and MSA with corresponding p-values.*

*Abbreviations: R: reading task, SP: sustained phonation task.*

### Speech tasks

For sustained vowel phonation, the subjects had to produce a sustained phonation of the vowel /a/ and /i/ for as long and steadily as possible. For alternating/sequential motion rates, subjects were instructed to repeat the syllables as accurately and quickly as possible within one breath. For text reading, subjects read the German linguistic text “Nordwind und Sonne” (4) at a comfortable loudness and speed.

### Acoustic analysis

The sound intervals used for formant measurements were chosen manually by visual inspection in Praat. Furthermore, all acoustic measurements were visually inspected in Praat and parameters were individually adjusted for each patient if the default parameters performed insufficiently. The default parameters used for the Praat routines are listed below. *Intensity variability*, *F0*, *F0 variability*, *HNR*, *jitter* and *shimmer* were calculated after removing the first and last 10% of the sustained phonation recording as recommended (2). Furthermore, *intensity variability*, *F0* and *F0 variability* were calculated after removing pauses from the text reading recordings (3). VOTs were measured based on visual inspection of raw waveform and wide-band spectrogram from the initial stop burst to the onset of periodicity associated with the vowel (5).

### Default Praat parameters

The complete Praat scripts including all parameters can be obtained from our github repository: github.com/t-haehnel/MSA-Speech-Analysis-Praat.

In general, we used sex-specific settings for pitch and formants:

**male:**

- minimum pitch: 60 Hz
- maximum pitch: 300 Hz
- formant ceiling: 4000 Hz

**female:**

- minimum pitch: 100 Hz
- maximum pitch: 500 Hz
- formant ceiling: 6000 Hz

The following parameters were used for **both** female and male:

- number of formants: 4
- formant window: 0.04 s
- minimum silence time for pause detection: 60 ms

All results were checked by visual inspection in Praat and default parameters were adjusted if needed.
